# Seroreactivity to SARS-CoV-2 in individuals attending a university campus in Bogotá Colombia

**DOI:** 10.1101/2021.03.15.21253609

**Authors:** John M. Gonzalez, Juan Carlos Santos-Barbosa, Catherine Jaller, German Otalora, Luis J. Hernandez, Marcela Guevara-Suarez, Silvia Restrepo

**Affiliations:** Grupo de Ciencias Básicas Médicas, School of Medicine, Universidad de Los Andes, Bogotá, Colombia; Medical Department, Universidad de Los Andes, Bogotá, Colombia; School of Medicine, Universidad de Los Andes, Bogotá, Colombia; Applied Genomics Research Group, Vice-president for research and Creation, Universidad de Los Andes, Bogotá, Colombia; Laboratory of Mycology and Phytopathology - Laboratorio de Micología y Fitopatología (LAMFU), Department of Chemical and Food Engineering

**Keywords:** Coronavirus, SARS-CoV-2, Covid-19, Serology

## Abstract

Most community-specific serological surveys for SARS-CoV-2 antibodies have been performed in healthcare workers and institutions. In this study, IgG antibodies specific to the virus were evaluated in individuals working at a university campus in Bogotá, Colombia. The aim of this work was to determine previous exposure to SARS-CoV-2 in those attending the campus during city lockdown. A total of 237 individuals, including 93 women and 144 men were evaluated using chemiluminescent detection of IgG anti N-viral protein between November and December 2020. There were 32 positives individuals corresponding to a seroprevalence of 13.5% (10 women and 22 men) and mostly asymptomatic (68.75%) and three cluster of seropositive individuals were identified. Only 13 of the seropositive individuals had previous positive detection of SARS-CoV-2 RNA by RT-qPCR performed in average 91 days before serological test. Seropositive individuals did not come from boroughs having higher percentages of SARS-CoV-2 cases in the city. This survey was carried out after the first peak of SARS-CoV-2 transmission in the city, and before the preparedness to reopening the campus for students in 2021, demonstrating a low seroprevalence in high percentage of asymptomatic. These results will help to evaluate some of the strategies stablished to control virus spread in the campus or other similar communities.

## Introduction

Covid-19 caused by SARS-CoV-2 emerged from China in 2019 (1). Due to the pandemic by the dissemination of the virus, several control and restricted community-based measures have been adopted worldwide (2). Coronavirus is transmitted directly from person to person or by contamination with fluids from infected individual; wearing face masks and hand washing are the most commonly used community personal protective measures to avoid infection spread (3, 4).

The recommended test during the acute infection is the nucleic acid amplification test (NAAT) using reverse transcriptase polymerase chain reaction (RT-qPCR) for the detection of viral ARN in secretions from the respiratory tract, mainly nasopharyngeal and oropharyngeal swabs (5). Serological tests are used to determine previous exposure to microbial agents; these are usually faster, cheaper, and some can be used as point-of-care (POC). Serological assays to measure SARS-CoV-2 specific antibodies are useful for surveillance studies and to determine the rate of exposure (6). There are different methods available including lateral flow immunochromatography (LFI), ELISA and chemiluminescence (CLIA) with a wide brand offer (7). Among other variables, serological tests can differ from antigens used for antibody detection and their performance characteristic such as sensibility and specificity (6, 8). A recent study has shown that the humoral immune response against SARS-CoV-2 is driven primarily by the spike (S) and nucleoprotein (N) viral proteins (9).

IgA and IgM specific antibodies can appear in average at 5 days post-infection (pi) and IgG can be detected at day 14 pi (10), and most people have already seroconverted between day 15 to 21 pi (10, 11). IgG specific antibodies against N or S protein can be detected as far as eight months after acute infection (12).

Most of epidemiological and serological specific population studies has been conducted in high-risk communities such as health workers and institutions (13, 14). Reopening of universityś campuses represent a challenge due to several factors including population commuting from different places, people socialization and gatherings, indoors activities such as lectures, laboratories and workshops that will help virus spread. The goal of this study was to assess the SARS-CoV-2 seroprevalence in individuals attending Universidad de Los Andes campus in Bogotá-Colombia, before reopening 2021 after the first peak of SARS-CoV-2 transmission in the city during 2020.

## Material and methods

### Type of study and population

This a transversal and descriptive study. Protocol and informed consent were approved by Ethical Committee of Universidad de Los Andes (Act No 1192-2020). The university is located in Bogota, Colombia, and has a population of 15,581 students, 1,808 teaching staff and 2,333 employees. (15). Volunteers of this study were working at the campus during city lockdown, when teaching was mostly remote. Individuals working in place were categorized and characterized according to age and presence of co-morbidities; they did not present any individual risk for complication due to SARS-CoV-2 or neither the members of the household. The risk characterization of individuals was based on the regulations of the Colombian health authority (resolution 666 and communication 30 of 2020).

### Blood sampling

Sampling was carried between the last week of November and first week of December 2020. Blood samples were drawn from the antecubital vein using a vacutainer without anticoagulant (BD, Franklin Lakes, NJ, USA). Blood was centrifugated at 2,300 rpm for 5 min, sera were separated and stored at 4ºC until used for antibodies detection the next day.

### Anti-IgG SARS-CoV-2 antibody detection

The chemiluminescence Abbott IgG Architect SARS-CoV-2 Assay (Abbott, Abbott Park IL, USA) was used. Test was run according to the manufacturers’ instructions. The antigen used for antibody detection is the nucleocapsid protein (N) from SARS-CoV-2. For interpretation, the index value reported as >1.40 was considered positive.

### SARS-CoV-2 detection by molecular biology

Nasopharyngeal swabs were collected in universal transport medium, the specimens were obtained from individuals attending the campus under the programs “Comunidad Segura” (Safe Community: epidemiological surveillance system for Covid-19 at the university) and COVIDA (Free SARS-CoV-2 testing project). Automated extractors, Genolution Nextractor® NX-48S (Seoul, Korea) with its nucleic acid reagents and Hamilton MicroLab Starlet MagEx STARline (Washington DC, USA), with the Quick-DNA/RNA Viral MagBead extraction Kit from Zymo Research (Orange, CA, USA) were used for the purification of RNA. RT-qPCR test was performed to determine the presence of SARS-CoV-2 RNA using U-TOP COVID-19 detection Kit that targets two regions SARS-CoV-2: ORF-1ab and N (SeaSun Biomaterial Inc., Daejeon, South Korea) and RNAse P as the internal control gene. This assay was run according to the manufacturerś instruction.

### Sociodemographic characteristic of the population

We obtained data concerning age and gender, commuting locality or residence, and type of work at the university. Individuals were asked for the presence of symptoms or signs associated to Covid-19 and contact with a SARS-CoV-2 positive person in the last week.

### Data presentation and analysis

Data is presented as percentages, mean or median and their respective standard (SD) deviation or interquartile ranges (IQR). A normality test (Shapiro-Wilk) was used for quantitative variables. Groups were comparing by non-parametric (U Mann-Whitney) or parametric (t-student) tests. Statistical analysis was done using the program PAST version 2.17c (http://priede.bf.lu.lv/ftp/pub/TIS/datu_analiize/PAST/2.17c/download.html)

## Results

A total 237 individuals were included in the study. Individual’s average age was 36.14 years (± SD 9.66), 93 participants (30.44%) were females. Average age was 33.8 years (± SD 7.97) for females and 38.06 years (± SD 9.83) for males (Table 1). No statistically significant difference was observed (p=0.17). Individuals (No. 214) live in 19 out 20 localities of Bogotá DC, except one (Sumapaz) placed in the rural area; the remaining individuals (no. 23) came from metropolitan municipalities (See Figure 1 and Table 2). According to activities at the university, 96 (40.5%) were in laboratories and workshops (includes post-grad students), administrative offices 81 (34.2%), general services 19 (8.0%), security 24 (10.1%), teaching staff 11 (4.6%) and health services 6 (2.5%). Most common reported symptoms at sampling were headache 3.3%, muscle pain 2.9%, coughing and nasal congestion 2.1%; no fever or dyspnea were reported. A total of 7 individuals reported contact with someone having symptoms related to Covid-19 within the week before sampling. IgG specific antibodies for SARS-CoV-2 N protein were detected in 32 individuals (13.5%) 10 women and 22 men (Table 1). This group presented a very low number of respiratory symptoms including: 2 nasal congestion, 1 coughing and 1 anosmia; no dyspnea was reported. Out of 32 individuals with positive antibodies, 13 work in the security department (40.6%), 11 in the laboratories (34.4%), 6 in administrative services (18.1%), 1 in general services (3.1%), and 1 in health services (3.1%). According to localities, seropositive came from 8 out 20, mostly from Rafael Uribe Uribe 6 (18%), Usaquén 5 (15.6%), San Cristóbal 4 (12.5%) and 4 (12.5%) outside Bogota (See map, Figure 1 and Table 2).

**Table 1.**
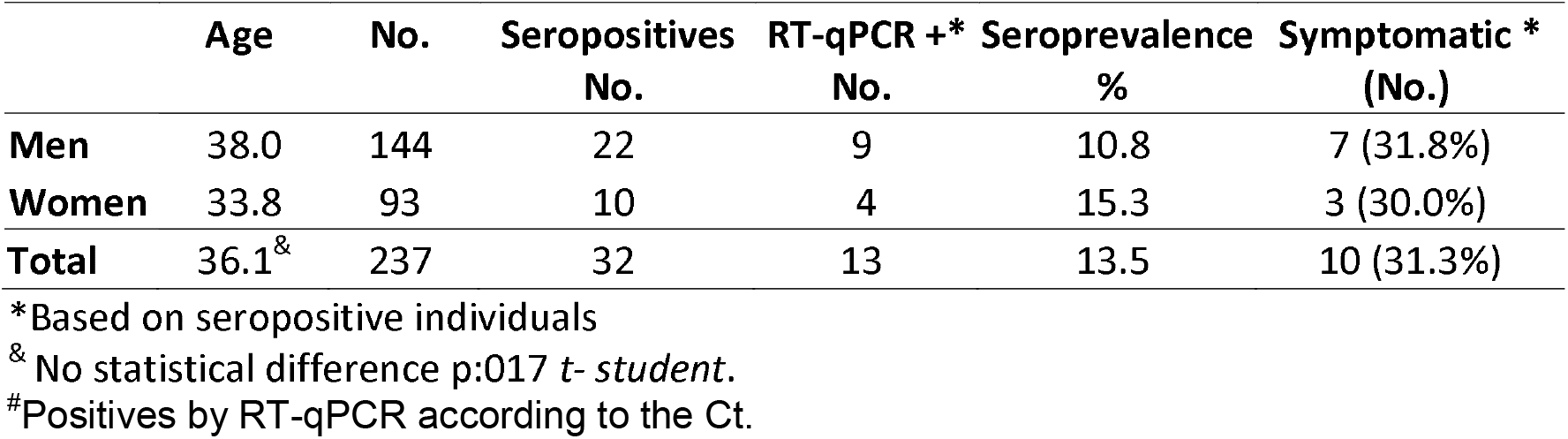
Summary of results.

**Figure 1.**
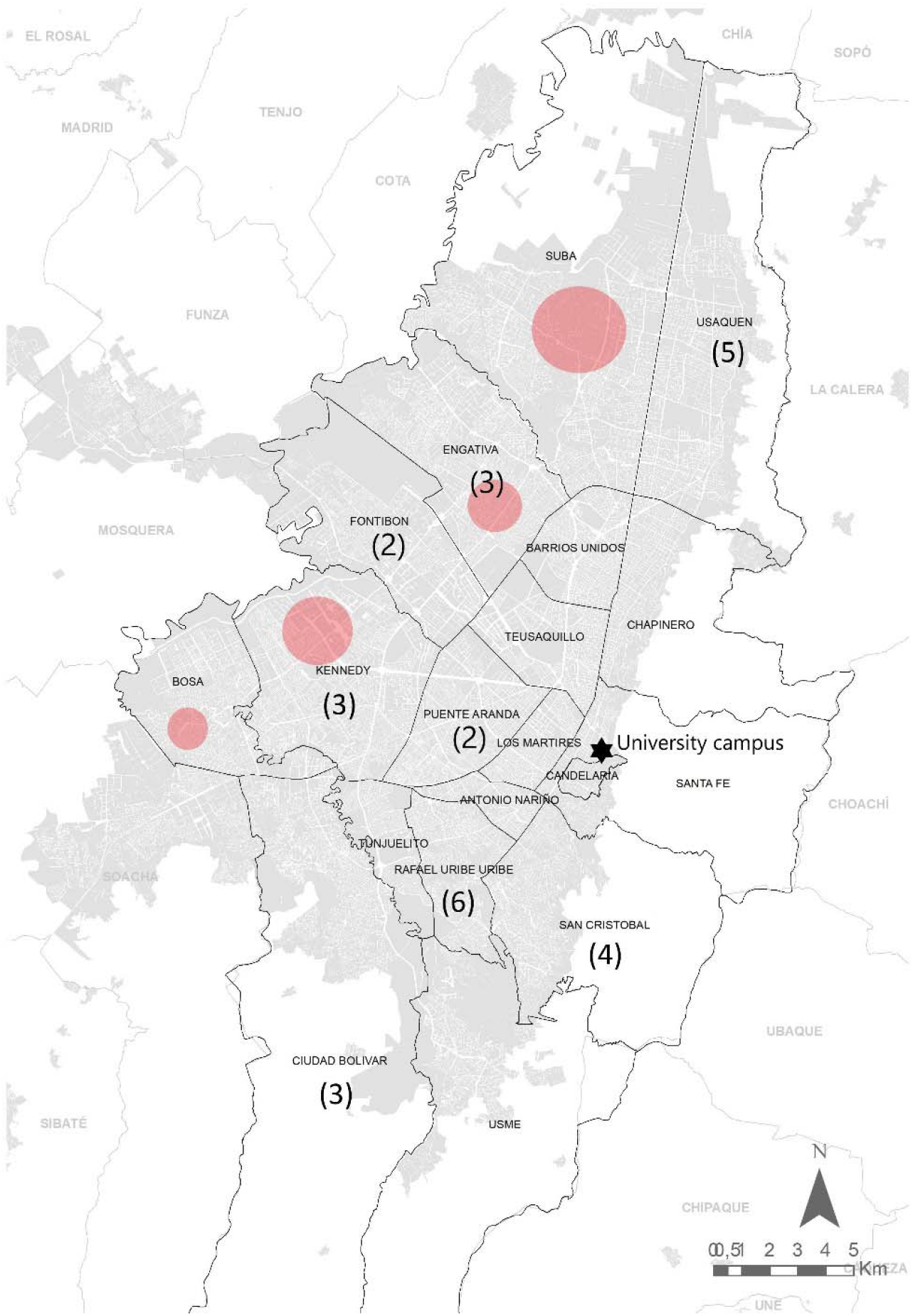
Localities in Bogotá where SARS-CoV-2 seropositive individuals lived. Number in parenthesis indicate the seropositive cases among university employees in each locality where they live. Red dots show the localities with higher number of SARS-CoV-2 positive individuals in the city. Map gently provide and adapted from José David Pinzon COVIDA program, Universidad de Los Andes.

**Table 2.**
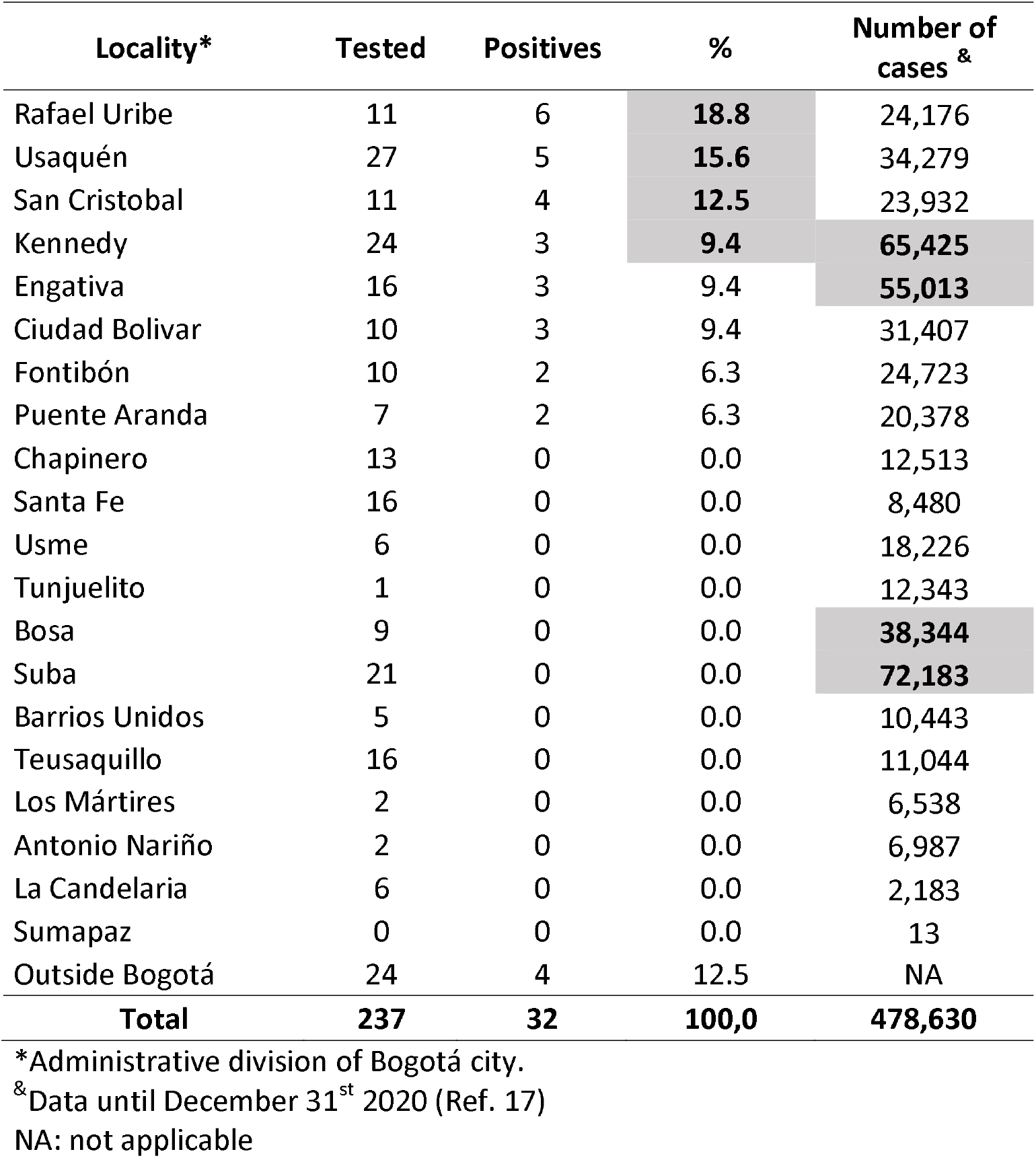
Comparison of seropositive individuals at the university campus and SARS-CoV-2 cases by localities in Bogotá.

| **Locality\*** | **Tested** | **Positives** | **%** | **Number of cases<sup>&</sup>** |
| --- | --- | --- | --- | --- |
| Rafael Uribe | 11 | 6 | **18.8** | 24,176 |
| Usaquén | 27 | 5 | **15.6** | 34,279 |
| San Cristobal | 11 | 4 | **12.5** | 23,932 |
| Kennedy | 24 | 3 | **9.4** | **65,425** |
| Engativa | 16 | 3 | 9.4 | **55,013** |
| Ciudad Bolivar | 10 | 3 | 9.4 | 31,407 |
| Fontibón | 10 | 2 | 6.3 | 24,723 |
| Puente Aranda | 7 | 2 | 6.3 | 20,378 |
| Chapinero | 13 | 0 | 0.0 | 12,513 |
| Santa Fe | 16 | 0 | 0.0 | 8,480 |
| Usme | 6 | 0 | 0.0 | 18,226 |
| Tunjuelito | 1 | 0 | 0.0 | 12,343 |
| Bosa | 9 | 0 | 0.0 | **38,344** |
| Suba | 21 | 0 | 0.0 | **72,183** |
| Barrios Unidos | 5 | 0 | 0.0 | 10,443 |
| Teusaquillo | 16 | 0 | 0.0 | 11,044 |
| Los Mártires | 2 | 0 | 0.0 | 6,538 |
| Antonio Nariño | 2 | 0 | 0.0 | 6,987 |
| La Candelaria | 6 | 0 | 0.0 | 2,183 |
| Sumapaz | 0 | 0 | 0.0 | 13 |
| Outside Bogotá | 24 | 4 | 12.5 | NA |
| **Total** | **237** | **32** | **100,0** | **478,630** |

\*Administrative division of Bogotá city.
<sup>&</sup>Data until December 31<sup>st</sup> 2020 (Ref. 17)
NA: not applicable

Out of 32 individuals with reactive serology, there were 13 individuals with RT-qPCR positive for SARS-CoV-2 RNA including 4 women and 9 men; 10 out of 13 were symptomatic at the time of the molecular assay. The RT-qPCR was done on average 91 days before the serological assay. One additional individual with positive RT-qPCR did not had detectable specific antibodies by CLIA. From 19 (6 women and 13 men) seropositive with previous negative RT-qPCR, done routinely as part of the surveillance program, none described symptoms associated with Covid-19 and only one endorsed previous contact with a SARS-CoV-2 positive person. In general, 31.25 % of the individuals that have positive serology were symptomatic (Table 1). Cluster defined as five or more epidemiological related cases were found in employees from security staff, engineering and sciences labs

## Discussion

The first imported case of SARS-CoV-2 infection in Colombia was reported in Bogotá on march 6^th^ 2020 (16); and until December 31^st^ 2020 the city reported 478,630 cases and a lethality of 2.1% (17). Bogotá entered in a preventive lockdown on march 20^th^, the first peak of infection occurs during July and August, and then the city reopened; at the same time employees and some postgrad students with lower risk returned to the campus with biosecurity protocols after September 1^st^. At this time, an effective reproductive number R(t) of 0.81 and around the sampling of this study R(t) averaged 1.16 (17). This opening brought a new challenge not only for the scholar community, but also for the people and business around it. Preparedness for possible reopening for the next academic cycle during early 2021 includes, among other protocols and guidelines, a seroprevalence assay conducted with individuals attending the university campus. People was characterized, and only those under 65 years old and without personal or familiar risk factors were allowed to attend. Most of the individuals performed activities in labs, workshops and administrative offices.

Our study evaluated presence of IgG anti-N protein of SARS-Co-2 by CLIA (18) on 237 individuals, from almost 300 people attending the campus in working days. Interestingly, most of the cases at the university did not come from localities in Bogotá with higher percentages of infection (Suba, Kennedy, Engativá and Bosa) (17). Seroprevalence among individual attending at Universidad de Los Andes in Bogotá was 13.5% which differs from other studies done in Colombia. In the national seroprevalence survey done by the Colombian National Institute of health (INS), crude and adjusted seroprevalence for Bogotá during October and November 2020 among 4,597 individuals was 26.3% and 30% a, respectively (19). Higher seroprevalence has been described for cities in the Caribbean area of Colombia reaching up to 55.3% (20). In a study carried in a university hospital in Bogotá during the first peak showed a baseline seroprevalence of 2.28% among health workers, increasing to 5.98% after 2-4 weeks of follow; with 38% of individual being pre-symptomatic or asymptomatic (21). Initial seroprevalence studies for SARS-CoV-2 IgG antibodies in Wuhan form March to April using a CLIA assay was from 3.2% to 3.8% showing information about the early spread and impact of the infection (22); similar values were found in systematic review worldwide until November 2020 showing 3.2% (IQR 1.0-6.4%) seroprevalence in general population (23). In this study near 70% of seropositive individuals did not recall any symptoms before the serological assay. One study showed that during SARS-CoV-2 infection the proportion of individuals without symptoms but positive for RT-PCR was 65.9% at the time of sampling, and 41.2% when using serological tests (24).

Reopening of colleges in Wisconsin (USA) showed increased in SARS-CoV-2 transmission among students demonstrating clusters of infection in three institutions; and virus sequencing showed rapid dissemination to the community (25). Congregation of students on and off campus helped to spread infection in a university campus in North Carolina (26). The close contact of young people at the campus, even if they are the less susceptible, can also facilitate the spread of infection to the university staff and their households. Therefore, cluster outbreaks and rapid dissemination are the two ways of transmission described in colleges and universities (26, 27). Here three cluster were identified, the largest being in the security staff, that have several epidemiological contacts due to their activities. The other two clusters were laboratories staff and postgrad student gatherings.

Different measures are adopted by the university with the aim to avoid spreading of the infection in the campus including characterization of individuals, online biosafety basic training and global screening using RT-qPCR for both symptomatic and asymptomatic employees. Also, mandatory university guidelines, including daily symptoms check through cellphone App (SeneCare), handwashing, use of face masks, physical distance and gathering limitations have been implemented. This study presented some limitation such as: number of samples, non serology comparison with other viral antigens and a retrospective analysis of RT-qPCR data, that did not include all the individuals. Although our data suggests that a very low percentage of the individuals have been infected, despite the fact of the use of public transportation and daily commuting between boroughs in the city with higher prevalence of infection. Although, this is a descriptive study, it was also found a high percentage of asymptomatic individuals and detection of clusters that indirectly helped to take some control measurements. Seroprevalence studies could help better understand the dynamics of virus transmission in university population as a subgroup of the broader general community setting or similar communities such as commercial or industrial.

## Data Availability

Data does not show in the paper will be available after request

## Acknowledgments

The authors want to thanks Abbott Colombia for the partial support of this work, and to the personal of the university health service for helping us with the blood sampling: Angelica González MD, María Imelda Hernandez and Giovanni Marín.

